# Titanium Fasteners In Endoscopic Mitral Valve Surgery

**DOI:** 10.1101/2020.06.03.20121400

**Authors:** Rafik Margaryan, Giacomo Bianchi, Tommaso Gasbarri, Giovanni Concistre, Marco Solinas

## Abstract

**Objective:** We sought to review our experience of new titanium knot fastener devices. We hypothesized that it might reduce the cardiop-polmonary bypass time, aortic cross-clamping time and intervention time.

**Materials:** We reviewed our electronic records in order identify the patients who underwent mitral valve (MV) repair and replacement in totally endoscopic setup. Surgical approach was trough limited right periareolar or inframmamary thoracotomy with mainly femoro-femoral arterio-venous cannulation. A part of patients underwent interventions using fast knotting system (FK group, Cor-Knot Device, ISL Solutions Inc) and remaining patients served as control group (conventional hand knotting, HK). We identified the FK patients and performed propensity score matching to match 1:1 ratio from main cohort using FK versus HK.

**Results:** A total of 306 patients underwent mitral valve repair or replacement on via right thoracotomy, 265 (87%) patients underwent using FK, remaining. There were on average 2.6 minutes of CPB time reduction (p = 0.64), and 3.1 minutes of CXC time reduction (p = 0.47). However, when dividing into procedures based on complexity, there were on average 8.6 minutes of CPB time reduction (p = 0.18), and 6.9 minutes of CXC time reduction (p = 0.16) in simple cases; on average in complex cases 12 minutes of CPB time was augmented (p = 0.24), and 2.5 minutes of CXC time was augmented(p = 0.76).

In propensity matched population the effect of CPB and CXC reduction was consistent and repeated and there were on average 0.5 minutes of CPB time reduction (p = 0.12), and 3.6 minutes of CXC time augmentation (p = 0.05). However, when dividing into procedures based on complexity, there were on average 0.2 minutes of CPB time reduction (p = 0.16), and 2.7 minutes of CXC time augmentation (p = 0.06) in simple cases; on average 5 minutes of CPB time augmentation (p = 0.34), and 14.2 minutes of CXC time augmentation (p = 0.58) in complex cases.

**Conclusions:** Titanium fasteners are useful tool to have in minimally invasive approaches, especially in complex cases and redo interventions. Titanium are comfortable and fast in many cases then conventional knot tying, but it is also expensive the traditional knotting. The titanium fasteners do not offer significant time reduction. In matched group the pattern of time saving were identical to main cohort.

## Introduction

Mitral valve dysfunction is the second-most common form of valvular defect in adults (Enriquez-Sarano, Akins, and Vahanian 2009). Last 15 years, minimally invasive mitral valve surgery (MIMVS) via right anterolateral mini-thoracotomy access has emerged and actually is an accepted approach for the management of mitral valve disease alone or in combination with tricuspid valve disease as a primiry approach or even in redo cases (Glauber et al. 2009; Murzi et al. 2014). This strategy originally was developed to decrease surgical trauma by minimizing the size of incisions, but as a metter of fact if offers more then just ‘cosmetic satisfaction’. It permits excellent exposure of the mitral valve, thereby avoiding conventional full sternotomy or resternotomy. This results in better visualization of the valve by the operating surgeon and, if video assisted, for other members (teaching involved), faster mobilization of the patient, faster healing, less pain, less infection and better cosmesis (Glauber et al. 2009, 2015). Fast tying can reduce CPB times and as a consequence reduce mortality and mobidity. The Cor-Knot automated fastener (LSI SOLUTIONS, Victor, NY, USA) is a alternative solution to knot pusher in limited access approaches or when CPB or cross-clamping time is cruscial, for example mitral valve surgery on beating or fibrillating heart. Mitral valve surgery without aortic cross-clamping is and valid alternative to restrenotomy in redo cases and can be performed with reasonable outcomes (Ad et al. 2015).

We hypothesized that there can be three main reasons for titanium fasteners use:

1. Reduction of aortic cross clamping time.
2. Reduction of cardiopolmonary bypass time.

### Objectives

We sought to analyse our primary experience with new fast knoting system (Cor-Knot automated fastener,LSI SOLUTIONS, Victor, NY, USA).

## Methods

### Participants

We identified 306 patiens redo patients (thoracotomy and sternotomy) between 2015 and 2019 at our centre were eligible for the present study (n=306). Data were collected prospectively, all of the minimally invasive mitral valve operations in this series were performed through the right minithoracotomy as described previously(Glauber et al. 2009, 2015). The primary outcomes for this study included myocardial ischemic time reduction (aortic cross-clamping time), and as secondary outcome cardiopulmonary bypass time reduction. For propensity score matching we have used comorbidity variables and demographic variables.

#### Surgical Techniques

We have used standard institutional techniques as described before(Glauber et al. 2009; Margaryan et al. 2019). Breifly, all patients were operated using institutional standardized endoscopic approach with femoro-femoral cannulation and without-rib spreading.

### Mitral valve repaire

The left atrium was opened in the Sondergaard groove and mainly used USB or Estech atrial retractors (Estech, San Ramon, CA, USA or USB retractor, Belgium) positioned through the same intercostal space as the thoracotomy medially or laterally to right internal mammary artery. The mitral valve was inspected and if reparable following techniqeues were used:

1. annuloplasty
2. leaflet resection
3. neochordal implantation
4. comisural or any other leaflet closure
5. Alfiery technique

In case of repair failure the standardized mitral valve replacement with bioprosthesis or bioprostesis were perfomred.

#### Statistical methods

Missing values were imputated using multipla imputation techniqeues in order to reduce bias and increase statistical power[12]. Two sided statistics were performed with a significance level of 0.05. For all analysis R Statistical Computing Environment[13] were used with RStudio (RStudio (2016). RStudio: Integrated development environment for R (Version 0.99.1198) [Computer software]. Boston, MA. Retrieved May 20, 2016. Available from http://www.rstudio.org/)

## Results

### Participants

In this sample of patients (n = 306) the mean age was 62.0 ± 11.7 years, 94 (31%) were female and the mean logistic EuroSCORE risk score was 5.0 ± 4.9% for mortality. None of patients suffered preoperative neurological disfunction. 200 (66%) were in III-IV class of NYHA. There were no significant differences in preoperative and postoperative variables between two groups (see Table 1 and 3). We performed 27 isolated MV replacements where 13 (4%) with biological valves and 14 (5%) with meccanical valves; remaning patients underwent ring annuloplasty. A total of 265 (87%) patients underwent operation using FK. A total of 306 underwent mitral valve repair or replacement on fibrilationg or beating heart. A total of 265 (87%) patients underwent operation using FK. There were on average 2.6 minutes of CPB time reduction (p = 0.64), and 3.1 minutes of CXC time reduction (p = 0.47). Howerver, when dividing into procedures based on complexity, there were on average 8.6 minutes of CPB time reduction (p = 0.18), and 6.9 minutes of CXC time reduction (p = 0.16) in simple cases; on average −12 minutes of CPB time reduction (p = 0.24), and −2.5 minutes of CXC time reduction (p = 0.76).

**Table 1:** Cor Knot Use in Mitral Procedures

|  | No | Yes | p.overall |
| --- | --- | --- | --- |
|  | *N=41* | *N=265* |  |
| Complex Mitral Procedure | 0.17 (0.38) | 0.29 (0.46) | 0.065 |
| Mitral Valve Procedure: |  |  | 0.594 |
| repair | 38 (92.7%) | 236 (89.1%) |  |
| replacement | 3 (7.32%) | 29 (10.9%) |  |

**Table 2:** Demographic variable divided by titanium fasteners use

|  | No | Yes | p.overall |
| --- | --- | --- | --- |
|  | *N=41* | *N=261* |  |
| Age | 63.4 (13.6) | 61.8 (11.4) | 0.478 |
| Sex: |  |  | 0.924 |
| Female | 12 (29.3%) | 82 (31.4%) |  |
| Male | 29 (70.7%) | 179 (68.6%) |  |
| LVEF | 57.8 (6.89) | 60.6 (7.43) | 0.022 |
| Hypertension | 0.32 (0.47) | 0.38 (0.49) | 0.410 |
| Smoke: |  |  | 0.426 |
| Ex smoker | 7 (17.1%) | 32 (12.3%) |  |
| Currently smoking | 5 (12.2%) | 22 (8.43%) |  |
| No smoker | 29 (70.7%) | 207 (79.3%) |  |
| Diabetes | 0.07 (0.26) | 0.06 (0.24) | 0.787 |
| Logistic EuroSCORE | 6.36 (6.46) | 4.75 (4.53) | 0.130 |
| BMI | 25.1 (2.89) | 24.7 (3.70) | 0.450 |
| Preoperative Creatinine | 0.93 (0.21) | 0.88 (0.24) | 0.228 |
| Arterial Cannulation Site: |  |  | 0.045 |
| Ascending Aorta | 4 (10.0%) | 6 (2.32%) |  |
| Axillary Artery | 0 (0.00%) | 1 (0.39%) |  |
| Femoral Artery | 36 (90.0%) | 252 (97.3%) |  |
| Venous Cannulation Site: |  |  | 0.044 |
| Right Atrium | 4 (10.0%) | 6 (2.32%) |  |
| Bicaval | 1 (2.50%) | 2 (0.77%) |  |
| Femoral Vein | 34 (85.0%) | 245 (94.6%) |  |
| Femoral and Jugular Vains | 1 (2.50%) | 6 (2.32%) |  |

**Table 3:** Demographic variable divided by titanium fasteners use

|  | **No** | **Yes** | **p.overall** |
| --- | --- | --- | --- |
|  | *N=39* | *N=39* |  |
| Age | 63.8 (13.7) | 58.4 (12.4) | 0.075 |
| Sex: |  |  | 0.479 |
| Female | 12 (30.8%) | 16 (41.0%) |  |
| Male | 27 (69.2%) | 23 (59.0%) |  |
| LVEF | 57.6 (6.98) | 66.4 (6.55) | <0.001 |
| Weight | 75.8 (12.2) | 337 (1154) | 0.166 |
| Height | 174 (9.70) | 167 (28.3) | 0.137 |
| Complex Mitral<br>Procedure | 0.18 (0.39) | 0.82 (0.39) | <0.001 |
| Intervention Time | 212 (233) | 257 (52.8) | 0.241 |
| CPB | 147 (32.5) | 159 (34.4) | 0.119 |
| CXC | 95.1 (25.7) | 107 (25.1) | 0.049 |

**Table 4:** Intraoperative variable divided by titanium fasteners use

|  | 0 | 1 | p.overall |
| --- | --- | --- | --- |
|  | *N=40* | *N=257* |  |
| CPB | 147 (32.1) | 144 (31.1) | 0.635 |
| CXC | 94.8 (25.5) | 91.7 (22.9) | 0.474 |
| Postoperative transfusion: |  |  | 1.000 |
| No | 7 (100%) | 34 (94.4%) |  |
| Yes | 0 (0.00%) | 2 (5.56%) |  |
| Hospital_Mortality: No | 39 (100%) | 246 (100%) | . |

#### Propensity score matching analysis

In propensity matched population the effect of CPB and CXC reduction was consistenta and repeated and there were on average 0.5 minutes of CPB time reduction (p = 0.12), and 3.6 minutes of CXC time augmentation (p = 0.05). Howerver, when dividing into procedures based on complexity, there were on average 0.2 minutes of CPB time reduction (p = 0.16), and 2.7 minutes of CXC time augmentation (p = 0.06) in simple cases; on average 5 minutes of CPB time augmentation (p = 0.34), and 14.2 minutes of CXC time augmentation (p = 0.58).

## Discussion

Our collective experience in minithoracotomy for mitral valve surgery alone with aortic cross clamping includes more than 2000 patients. The very small sample of patines are addressed to minimal thoracotomy (endoscopic approach) using video guiadence. We demonstrate that time reductin in the main cohort is present, but it is not significant. In propensity matche group the patern was repeting itself, except in complex cases there were CPB and CXC time agumentation. It is due the unbalanced complexity patients percentate in the FK group (see Table 3) In our clinical practice minimally invasive mitral valve approaches with femoral cannulation and retrograde perfusion is prefered way for endoscopic mitral valve surgery because of limited thoracotomy and no rib retraction. In the intial echo to minimally invasive techniques for mitral valve surgery, some authors reported and expressed some concerns that the results of these approaches were inferior to those of median sternotomy, particularly with regard to stroke and other major complications (Vanermen et al. 1999; Mohr et al. 1998). Recent publications have demonstrated the safety and effectiveness of minimally invasive mitral surgery also with a fibrillatory arrest method(Umakanthan et al. 2008,@kilo_minimally_2013). Innovative aspect extends bounderies of the minimally invasive cardiac surgery; innovative products are very important for developing new approaches. Titanium fasteners have demonstrated to be d easy to use reduce the paravalvulare leaks(Lee et al. 2016). Plestis have demonstrated that titanium fasteners can decreas intraoperative times, blood derivated transfusion and it allowed to be more reproducible (Plestis et al. 2018). There are new reports that use titanium fasteners for left ventricular assist device implantation, which confirms this accessories utility(Johnson et al. 2018).

### Limitations

This study has several limitations: limited sample size, retrospective fashion of the study, bias of selection to be addressed in one or other approach, single center experience.

## Conclusions

Titanium fasteners are useful tool to have in minimally invasive approaches, especially in complex cases and redo interventions. Titanium are comfortable and fast in many cases then conventional knot tying, but it is also expensive the traditional knotting. The titanium fasteners do not offer significant time reduction. In matched group the pattern of time saving were identical to main cohort.

**Figure 1.**
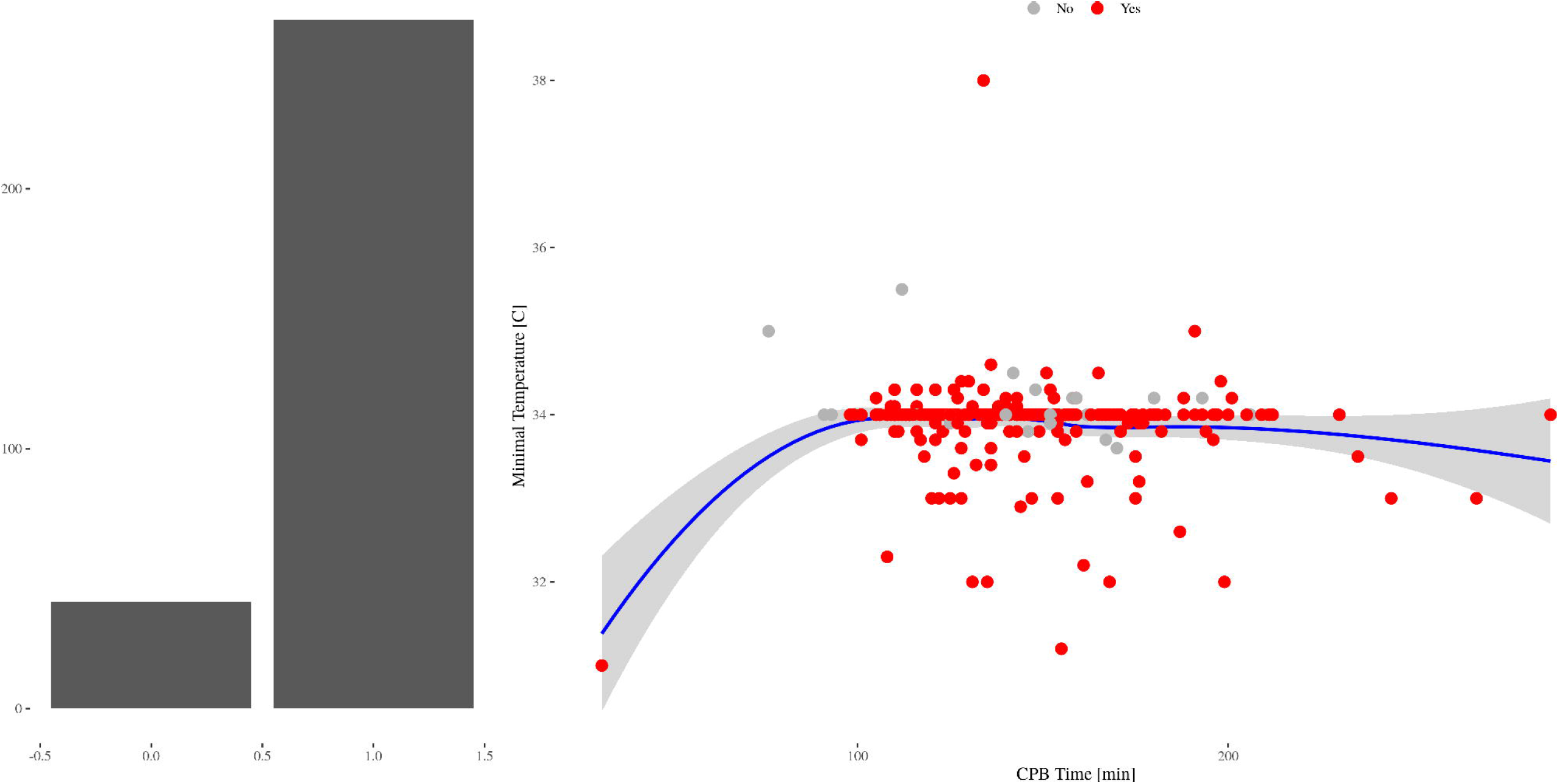
FK distribution; CPB and minimal temperature relationship depending on FK use.

## Data Availability

Some data will be made available.

